# Best Practices on Big Data Analytics to Address Sex-Specific Biases in our Understanding of the Etiology, Diagnosis and Prognosis of Diseases

**DOI:** 10.1101/2022.01.31.22270183

**Authors:** Su Golder, Karen O’Connor, Yunwen Wang, Robin Stevens, Graciela Gonzalez-Hernandez

**Affiliations:** Department of Health Sciences, University of York, York, UK; Department of Biostatistics, Epidemiology and Informatics (DBEI), University of Pennsylvania, Philadelphia, Pennsylvania, USA; Annenberg School for Communication and Journalism, University of Southern California, Los Angeles, USA

**Author notes:** email: [, ].

**Keywords:** machine learning, natural language processing, bias, health disparities, gender disparities, ethics

## Abstract

A bias in health research to favor understanding of diseases as they present in men can have a grave impact on the health of women. This paper reports on a conceptual review of the literature that used machine learning or NLP techniques to interrogate big data for identifying sex-specific health disparities. We searched Ovid MEDLINE, Embase, and PsycINFO in October 2021 using synonyms and indexing terms for (1) “women” or “men” or “sex,” (2) “big data” or “artificial intelligence” or “NLP”, and (3) “disparities” or “differences.” From 902 records, 22 studies met the inclusion criteria and were analyzed. Results demonstrate that the inclusion by sex is inconsistent and often unreported, although the inclusion of men in the included studies is disproportionately less than women. Even though AI and NLP techniques are widely applied in health research, few studies use them to take advantage of unstructured text to investigate sex-related differences or disparities. Researchers are increasingly aware of sex-based data bias, but the process towards correction is slow. We reflected on what would be the best practices on using big data analytics to address sex-specific biases in understanding the etiology, diagnosis, and prognosis of diseases.

## 1. INTRODUCTION

Recent advances in data science and digital epidemiology have unlocked an unprecedented amount of data for analysis, and uncovered previously unseen sex-specific patterns that point at marked differences in disease symptoms, progression, and care that disproportionately affect women of all ages. In 2016, the NIH published a guidance document (1) and changed its policy for reviewing proposals whereby accounting for “sex as a biological variable” became a required and scorable aspect of the research strategy, highlighting that “an over-reliance on male animals and cells may obscure understanding of key sex influences on health processes and outcomes.” The statement from the NIH only highlights the tip of the iceberg: biases at the start of the research pipeline impact our capacity to derive knowledge upstream compounding the problem even further as the same biases continue at different levels. Thus, in pre-clinical research, male mouse models are overall more represented than female models (2, 3, 4). The reason behind this practice is a concern that the reproductive cycle can induce variability in the experimental design and a historical presumption that sex does not matter, which continues to prevail despite the efforts of the NIH and evidence to the contrary in several fields (2, 5, 6). Sex differences exist in every tissue and every organ system in humans (7, 8). Further up, recruitment for clinical trials often avoids women of childbearing age due to safety concerns, particularly in phase I and phase II trials. Even when both sexes are included, sex-differences are often not reported. Consequently, some diagnoses and treatments are not adequately developed or evaluated in women.

Because of seemingly routine research decisions such as which cell lines or animal models to use, or even well-intentioned decisions such as protecting women in reproductive age, health research has had and continues to have a recognized intrinsic bias that favors knowledge and understanding of common diseases as they present in men. This can have a grave impact on the health of women. For example, the first scientific statement from the American Heart Association on acute myocardial infarction in women was published only in 2016, reporting on trends and knowledge gained over the prior 15 years. Before that, knowledge about how myocardial infarction presents in women (9), and the signs and symptoms specific to women were not widely known or publicized. As knowledge improved, there were marked reductions in cardiovascular disease mortality in women (see Figure 1) (10), demonstrating the devastating impact that failure to recognize early symptoms and other gaps in knowledge have on the affected population. Even despite recent advances, women are less frequently referred for appropriate treatment (9, 11). Indeed, the problems in cardiovascular disease may be mirrored in other disease areas. Sex bias is known to exist in respiratory diseases (12, 13, 14), neurological disease (15), diabetes, mental health disorders (16), cancer (17), autoimmunity (18), as well as physiological processes such as pain sensitivity (19) and aging of the brain (20).

**Figure 1.**
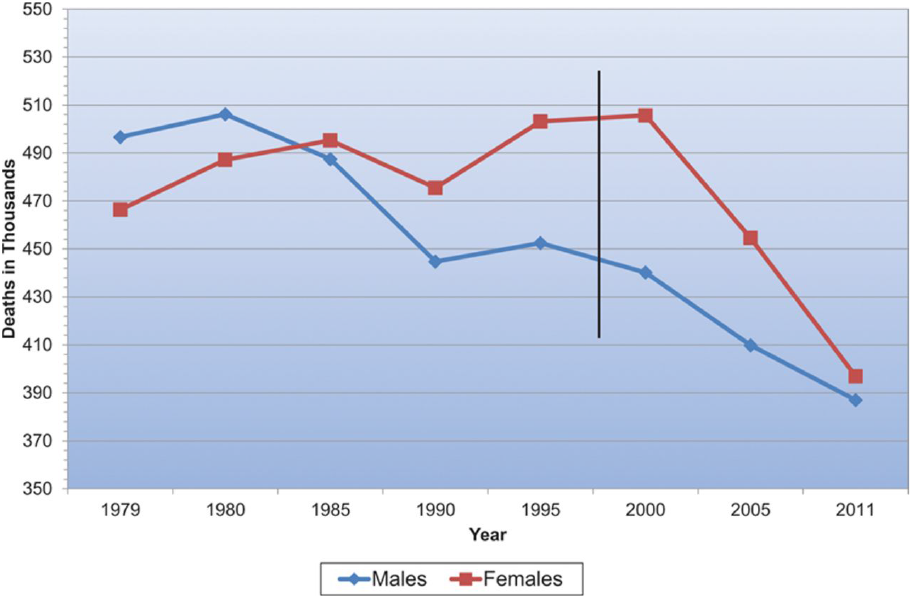
Cardiovascular disease mortality trends for men and women in the United States from 1979 to 2011. Reprinted from Mozaffarian et al. (2015). Copyright © 2015 American Heart Association, Inc.

The wide adoption of Electronic Health Records (EHRs) has resulted in large repositories of clinical and genomic data from a broad and diverse set of patients and has opened the door to studies at a scale never seen before. The use of such data for digital epidemiology could help alleviate the effects of the early biases and address the sex-specific knowledge gaps by providing readily available real-world data (RWD) for observational studies encompassing a diverse population to derive real-word evidence (RWE). The United States Congress addressed RWE in the 21st Century Cures Act of 2016. The Act required that the Food and Drug Administration create by 2018 a pathway to allow RWE to support new drug indication and post-marketing surveillance (21).

However, despite its wide use and availability for research, roughly 70%-80% of the data (22) in EHRs is never used, as most published research that uses such repositories limits the study to its structured portion (data in discrete fields, such as dates, ICD-10 codes, or test results, among others). The much more abundant and information-rich portions of the records (the unstructured free text in the admission, progress, and discharge notes, and radiology or pathology reports) are largely left untouched. Just as an example from the present study, in our MEDLINE search we found 67,424 papers as a result of our search of sex-specific keywords plus the keywords indicating ‘differences’ before adding the keywords of methods that could signal the use of unstructured data (such as ‘nlp’, ‘text mining’, ‘deep learning’ and others). Once those were added we ended up with only 469 papers from MEDLINE, which could indicate that unstructured data is likely utilized only in less than 1% of the studies. This could be due in part to the many challenges that using the unstructured data would present, including its inherent ambiguity, and to the fact that the methods needed to extract and convert the free-text data are still developing and are not widely accessible to the average health care professional researcher, particularly in relation to longitudinal data. In addition, access to the unstructured portion of the records is more limited, and there are really not many agreed-upon ‘best practices’ to do so. The motivation to use the unstructured text for filling the gaps in knowledge is strong, however. Free text is generally preferred by clinicians (22), as it can reflect their thought process and is perceived as more accurate, reliable, and understandable (23), and allows them to express the complexities of diagnosis and interventions in clinical practice and the ambiguity of symptoms (such as something being “more or less resolved”, or happening “often over the last month”) in a way that is impossible with structured data (24).

Furthermore, careful design and systematic use of advanced Artificial Intelligence (AI) methods, such as Natural Language Processing (NLP) and Machine Learning (ML) could help address the gaps in knowledge. This was demonstrated in a recently published retrospective observational study using EHR data (25). The dataset included 10,840 clinical notes. In the study, a set of clinical concepts was extracted from EHR structured and unstructured data using traditional query techniques and artificial intelligence (AI) technologies. Performance was evaluated against manually annotated cohorts. Accuracy was compared to pre-defined criteria for regulatory grade. Individual concept occurrence ranged from 194 for coronary artery bypass graft to 4502 for diabetes mellitus. For the structured data, the average recall and precision were 51.7% and 98.3%, respectively and 95.5% and 95.3% for the unstructured data, respectively. Thus, for each clinical concept, accuracy for the strucutred data was below regulatory-grade, while the unstructured data met or exceeded the criteria, with the exception of medications. In another study (26) the authors sought to quantify the occurrence of serious and mild-to-moderate hypoglycemia using a large EHR database in the US, comparing estimates based only on structured data to those from structured data and natural language processing (NLP) of clinical notes, using 844,683 clinical records. They found that the use of NLP more than doubled the completeness of hypoglycemia capture overall relative to measures from structured data, and increased capture of non-serious events more than 20-fold.

The use of these methods can also allow the integration of data not traditionally used in clinical studies, such as the data in registries and regulatory data, or data that directly captures the patient perspective, such as what is posted in Twitter, Reddit, or health forums. Although social media text mining research for health applications is developing, the domain has seen a surge in interest in recent years. Numerous studies have been published of late in this realm, including studies on pharmacovigilance (27), medication non-adherence (28), toxicovigilance (29), foodborne illness (30), and tracking infectious/viral disease spread (31, 32). The onset and spread of SARS-COV-2 causing the COVID-19 pandemic has been the impetus for new health related social media text mining research to look at symptoms (33, 34, 35, 36, 37, 38) and potentially predict new outbreaks (38, 39, 40). However, methods to determine the biological sex of a social media user are limited and imprecise, and research using these alternative data has not yet reached the point where sex-specific health outcomes can be accurately elucidated. We only found two papers that attempted to do so (41, 52).

When speaking of women’s health, this study focuses on reported sex-specific differences in studies of human disease that generally refer to biological sex. Further work may be needed to identify gender disparities in health. Gender disparities are multifaceted and are related to the social status of women, as well as real and perceived biological differences and computational health research (CHR) can aid in identifying them. These methods can also be used to develop models that sidestep traditional gender biases, and develop models that can improve diagnosis, care and ultimately health outcomes for all women. Going one step further, the scale of data that can be used in CHR also allows for targeted examination of health outcomes among marginalized women. It is among Black, Indigenous, and Latina women where we can expect to find the most significant health disparities, but traditional health research is often underpowered to detect differential outcomes among them. Overall, computational health research is not bound by sample size limitations and can serve as a powerful tool to achieve equitable care for all women.

## 2. METHODS

In this paper, we aim to capture an understanding of how methods have been developed that interrogate big data to identify sex-specific disparities in health, and reflect on what would be the best practices for the purpose. To this end, we conducted a conceptual review of the literature. We had a broad understanding of the concepts or themes we anticipated to provide our framework and we anticipated that the literature would be too diverse to synthesize in a traditional systematic review.

The first stage of our review was to structure the question. We used a PICO (Population, Intervention, Comparator and Outcome) format to structure the question and thus inform our inclusion and exclusion criteria and search strategy. In terms of the population (P) we included both males and females of any age with any disease or condition. Studies limited to just men and studies limited to just women were excluded. We also excluded all animal studies.

While within our question, there is no traditional intervention (I) as such we considered any paper that had used artificial intelligence on any big data (structured or unstructured). We were particularly interested in machine learning (ML) or natural language processing (NLP). We excluded studies that only investigated using AI on images or videos, or genomic data. We also excluded studies that conducted only manual analysis of data or did not utilize any AI methods. We also excluded those that used AI solely to infer or predict sex or gender.

As for the comparator (C), all studies were required to compare males to females (or vice versa), as we are interested in sex-specific differences. With regard to the outcome(s) (O), we included any measurement of health disparities or differential health data by sex. We excluded those studies limited to other demographic data health disparities, such as race, social class or geographical region, or that simply expressed a sex difference as a demographic characteristic of its study population without further analysis of its impact on outcomes.

We searched Ovid MEDLINE, Embase, and PsycINFO on the 19th October 2021. Our search strategy was broad, incorporating synonyms and indexing terms for women or men or sex (representing the population of the study (P) or the comparator (C)), big data or artificial intelligence (AI) (representing the intervention (I)), and disparities or differences (representing the outcome(s) (O)).

We anticipated, however, that the study of sex-specific disparities is likely to be a secondary or tertiary focus of many studies. Such studies will be difficult to identify given that gender is unlikely to be mentioned in the title, abstract and even indexing in bibliographical records in databases such as Medline and Embase. We therefore also conducted citation searches for all studies that met our inclusion criteria as well as reference checking of any related reviews and our included studies. Consequently, we acknowledge that the views and discussion of the current ‘state of the art’ in this area presented here may be limited by being unable to uncover all examples in the literature.

The results of our searches were imported into an Endnote Library and then sifted by title and abstract for potentially relevant studies. The full-text articles of any potentially relevant studies were then independently assessed by two reviewers to ascertain whether they did indeed meet our inclusion criteria.

We extracted the following data from the studies that met our inclusion criteria, the disease or condition studied, the purpose, data source and cohort size of the study, the gender distribution, the AI methods including features/predictions, annotation and validation, the results of the study and whether separate models were used for males and females and code made available. We then summarized the included studies according to the attributes of the studies.

## 3. RESULTS

Our search returned 902 unique records (1331 records before deduplication) and after reviewing the abstracts, 62 references were included for screening of the full text. After review of the full text, 22 studies met our inclusion criteria and were included while 40 were excluded. The main reason for exclusion was the study did not use any machine learning or NLP techniques. Other reasons, in order of prevalence, were the reference retrieved was only an abstract or was not an original research article, the data used was exclusively from our excluded criteria, the reported ML study only classified users by gender, and the population did not meet our inclusion criteria.

We analyze next the reviewed studies based on their knowledge area, data sources, the application of AI methods, and the proportion of men and women in the study samples.

### 3.1. Knowledge Area

#### Prescription rates

These have been identified in women and men while controlling for confounding factors such as sociodemographic, clinical (e.g., psychiatric comorbidities and substance use), neutropenia, functional factors (e.g., problems with occupation), and clinical monitoring (43).

#### Symptom Presentation

Differences in symptom presentation at first episode psychosis (FEP) have been studied using NLP and ML in EHRs (44) and diagnosis and clinical manifestations of COVID (45) and tinnitus (46), Friedreich’s ataxia (FRDA) (47) and Alzheimer’s disease (48). Using the data output from medical devices, sex-specific differences were examined for gait kinematics in patients with knee osteoarthritis (49) and heart rate variability (50). The importance of gender balance in sample sets for identifying phenotypic characteristics for diagnosis has also been emphasized with a study on sex differences in autism (51). The presentation of mental illness disclosure on social media was assessed by sex (52).

#### Risk Predictors

Studies have differentiated the risk of suicide attempt and self-harm (53), metabolic syndrome (54) initiation of cannabis use (55), the risks of loneliness (64), obesity (56), musculoskeletal disorders (57) and opioid use post-treatment (58) using EHRs, as well as other datasets.

#### Treatment

Only two studies found sex-specific differences in treatment, specifically indicated in the treatment of COVID (45) and in treating the effects in tinnitus (46).

#### Prognosis

The study of disparities in prognosis is of particular importance. Sex-specific differences in survival were identified using machine learning algorithms in lung cancer patients (Wang 2021), and those undergoing cardiac surgery (60).

#### Comorbid Conditions

Sex-specific differences in multimorbidity networks were examined using a large set of EHR data (61).

### 3.2. Data Sources

The data sources for the studies were varied, with most utilizing information from EHRs or other clinical databases. Within these studies, the majority only included structured data in their models (47, 48, 49, 50, 58, 59, 60, 61, 62) while a few incorporated both structured and unstructured data from these resources (45, 43, 44, 63). Another source of data was surveys, questionnaires, or interviews, where again the majority of studies used only structured data in their models (46, 53, 54, 51, 57), with only one study applying NLP methods and used both structured and unstructured data (64). Two of the studies (52, 56) explored data retrieved from Twitter, using NLP methods to detect and extract relevant information and machine learning methods for classification of the tweets.

The majority of the included studies employed only machine learning techniques (15/22, 68.2%) to uncover gender disparities. These included traditional machine learning methods such as Support vector machine (SVM) (49, 50, 51), classification tree analysis including random forest (53, 58, 60), XGBoost (59) and C4.5 (47), regression models RIDGE (46) and logistic regression (59, 55) or other regression models, such as Multivariate adaptive regression splines (MARS) (57). Other studies used artificial neural networks, including self-organizing maps (SOM) (48, 61), and AutoCM (54). One study used topic modelling to identify disease disclosure (52). Two of the included studies (2/22, 9.1%) applied NLP techniques only to extract information from the free text of EHRs (44, 63). The remaining studies (5/22, 22.7%) used both NLP to identify and extract information and machine learning methods for classification or prediction (43, 41, 45, 52, 64).

### 3.3. The Application of AI Methods

The ways in which NLP or ML are applied vary considerably. Some studies use text mining or NLP to extract data from unstructured or free text from sources such as EHRs. This is the case for a study on the diagnosis and clinical management of COVID-19 using EHRs from Spain (45), a study on clinical presentation and illicit substance use during first episode psychosis in London, England (44), another study on reasons for discontinuation of lipid-lowering medications in patients with chronic kidney disease in the U.S. (63) and another on clozapine prescription in schizophrenia in London, England (43). In each of these cases statistical analysis was then conducted using the data extracted.

Most of the studies explored the role of sex-specific differences in the creation of classification or prediction models for a variety of conditions including, Alzheimer’s disease (48), autism(51), cancer (59), cardiovascular disease (47, 60, 63), musculoskeletal disorders (57), substance use (44,55, 58), mental illness (43, 52, 62, 64) obesity/physical activity (41, 54) and tinnitus (46). Within these studies, nine developed separate models for males and females finding these models were better able to discriminate sex-specific determinants of the trait of interest.

One study used machine learning (ML) methods to identify physiological differences between males and females that may be missed using traditional statistical techniques. This study examined sex differences in gait in a gender-matched study to identify differences between healthy and osteoarthritis patients (49). This study found complex relationships amongst variables that may not be discovered using inferential statistics and differences that were identified only using sex-specific comparisons (49). Another study examined sex-specific differences in heart rate variability from touch stimulus (50), noting that the SVM classifier achieved higher accuracy when the models were trained and deployed on gender specific cohorts.

Disease differences by sex were also studied to find the health disparities in disease diagnosis to understand multimorbidity networks (61). A summary of the studies included in the review can be found in Table 1 and the AI methods and results in Table 2.

**Table 1.** Summary of studies including data source, cohort size and gender distribution.

| Study | Disease/Condition | Data Source | Cohort Size | Gender Distribution |
| --- | --- | --- | --- | --- |
| Baron-Cohen et al. | Autism | Questionnaire | 715 | Cases:178 M, 217 F; Controls: 152 M, 168 F |
| Gradus et al. | Suicide risk | EHR | 279 286 | Cases: 10 152 M, 3951 F; Controls: 130 591 M, 134 593 F |
| Tokodi et al. | HF patients | EHR | 2,191 | 1637 M, 554 F |
| Busto Serrano et al. | Musculoskeletal disorder | Survey | 43 500 | 21 922 M; 21 578 F |
| Davis et al. | Post treatment opioid use | Structured clinical data | 1126 | 508 F, 618 M |
| Vigna et al. | Metabolic Syndrome | Interview/questionnaire | 210 | 68 M; 142F |
| Wang et al. | Lung cancer | SEER database | 28 458 | 13 458 M; 15 000 F |
| Spechler et al. | Drug and alcohol use | Survey, EHR | 1587 | Cannabis use: 207 M, 158 F; Comparator: 538 M, 678 F |
| Gradus et al. | Suicidal Ideation | Survey | 2238 | 1062 M; 1099 F |
| Ghorbani et al. | FRDA | Structured clinical data | 600+ | Not reported |
| Cesare et al. | Physical activity level | Twitter | 481 146 | 267 758 M; 213 388 F |
| Badal et al. | Loneliness | Interviews/assessments | 88 | 29 M; 51 F |
| Niemann et al. | Tinnitus | questionnaire | 1628 | 800 M; 828 F |
| Grossi et al. | Alzheimer's Disease | Structured clinical data | 211 | 68 M; 143 F |
| De Choudhury et al. | Mental health | Twitter | 1 208 727 | 133 608 M, 192 265 F |
| Ancochea, et al. | COVID-19 | EHR free text | 4780 | 2337 M; 2443 F |
| Wesley et al. | Schizophrenia | EHR structured and free text | 2244 | 1441 M; 803 F |
| Irving et al. | Substance use | EHR structured and free text | 3350 | 2092 M; 1258 F |
| Morrison et al. | Chronic kidney disease | EHR structured and free text | 14 034 | 5675 M; 8359 F |
| Kalgotra et al. | multimorbidity | EHR structured data | 22.1 million | 9.9m M; 12m F |
| Nardelli et al. | Heart rate variability (HRV) | HRV series from ECG | 32 | 16 M; 16 F |
| Phinyomark et al. | Osteoarthritis of the knee | Structured clinical data | 143 | Cases: 45 M, 55 F; Controls: 18, M, 25 F |

**Table 2.**
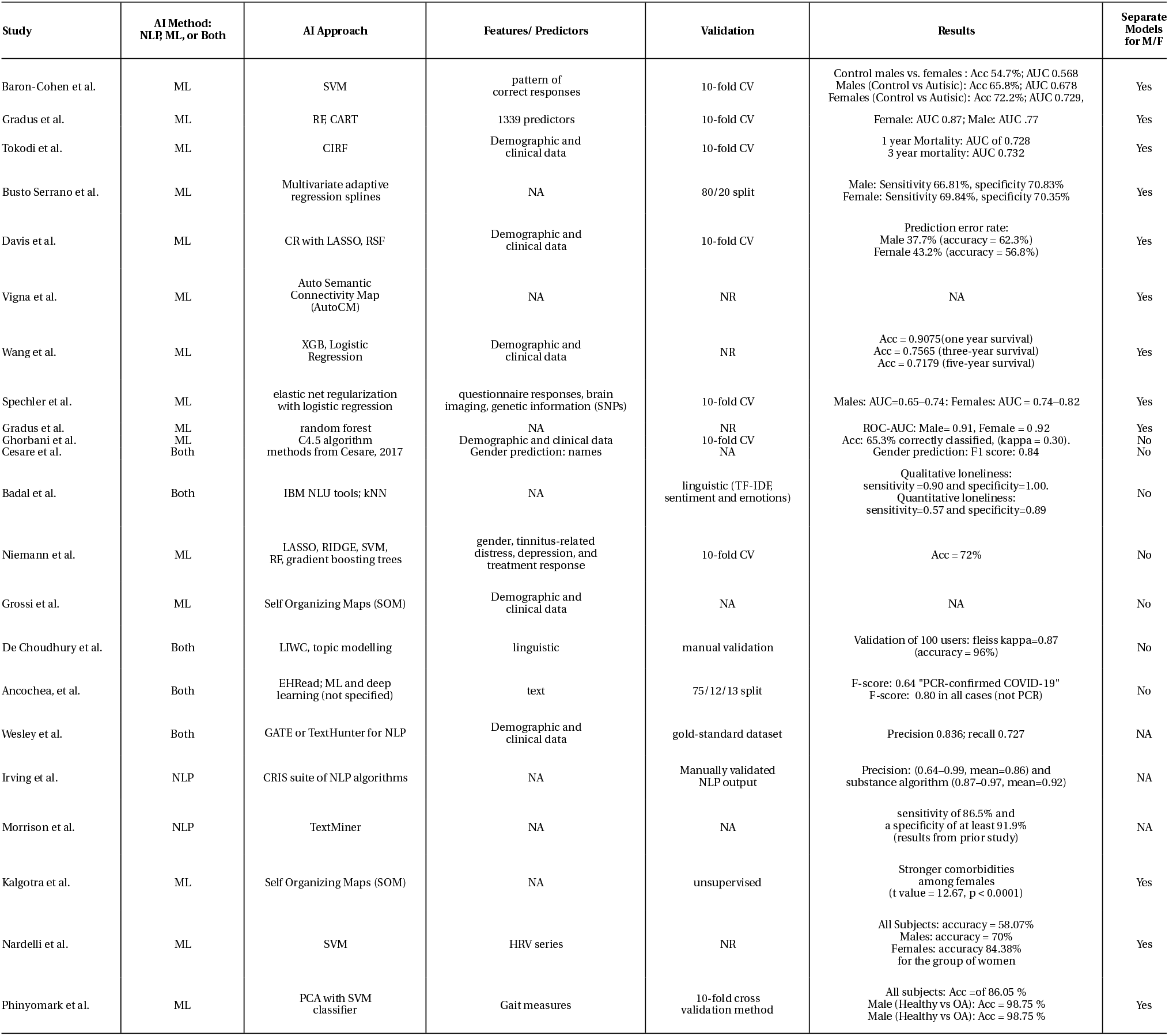
Summary of AI methods and results of included studies.

### 3.4. Study Samples

Among the papers reviewed, the proportion of women included varied significantly and in a two instances sex was not reported (47, 52). Most studies included more women than men (48, 54, 55, 61, 64), several more balanced samples (46, 59, 50, 53). In two studies (62, 55), which have a majority of male participants in the outcome group but the majority of participants female in the comparison group.

## 4. DISCUSSION

Sex and gender have a profound impact on the diagnosis, progression, care of illness and health outcomes among women (5). Identifying and addressing sex- and gender-related issues in health care is a crucial step towards achieving health equity (65). This research is a global priority named within the WHO’s Sustainable Development Goals, and a steppingstone to promoting human rights, and achieving health for all (66).

However, there is increasing recognition that the current attention paid to gender bias in health is suboptimal (67) and this appears to extend to computational health research. Our review demonstrates that while CHR, and AI specifically has been used to study a variety of aspects of health care, there have been relatively few cases where such techniques have been used to investigate sex-related differences or disparities. While researchers are becoming increasingly aware of male data bias, where data collected, analysed and used in decision making has a bias towards men (5), this process towards correction is slow. We can be careful not to repeat these mistakes i n computational health research. Future studies of big data from real-world settings, however, may help us to distinguish between sex differences and gender disparities driven by bias, sexism or gendered hierarchy. As some differences are explained by underlying biology, others are actually disparities that result from inadequate or inappropriate care, or differences in social-economic factors or lifestyle factors (such as alcohol consumption, diet, smoking, and exercise) (68, 69). CHR methods and big data can be useful tools to disentangle the two.

Artificial intelligence should be performed on large scale data to make stronger sex-specific inferences. We know that sex-specific differences exist but we need more detailed information to inform prevention, diagnosis, treatment and clinical management. Big data has the potential to decompose gender related heterogeneity leading to more personalised informed decision making in healthcare.

### 4.1. Potential Biases in Studies and Implications for Good Practices

Surprisingly, among the papers reviewed, the inclusion by sex is inconsistent and often not reported. In the majority of the studies, the inclusion of men is often disproportionately less than women (64, 48, 61, 55, 54), except for a few that have more balanced subgroups (53, 50, 46, 59). There are two exceptions (55, 62), which have a majority of male participants in the outcome group but the majority of participants female in the comparison group. In two other studies (52, 47), the distribution of males and females was not reported. The gender-skewed data distributions, if not transformed, may lead to spurious findings in gender-specific differences, therefore challenging the validity of the studies and resulting in biases in generating understanding of the etiology, diagnosis and prognosis of diseases. For example, autism research normally involves male dominated study samples; (51) used a sex-balanced sample of men and women with Autism and found the absence of typical sex difference in eye test. As (49) postulate, the lack of consensus in previous studies with respect to certain diseases could be due to the results of mixed-gender cohorts. Thus, it may be a good practice to generate gender-specific models. Moreover, gender is undoubtedly confounded with other variables such as age in the etiology of diseases (49). When creating sub-groups for research purposes, researchers should inspect gender systematically in relation to other variables rather than as considering it a standalone factor.

### 4.2. Next Steps

Thoughtful use of big data can aid our understanding of women’s health needs. Automated methods can be deployed to identify gender-based disparities in diagnosis and care (66). This is a powerful tool to achieve equity in diagnosis, care and health outcomes for women, across a variety of health domains.

Automated methods should also be used to investigate the interplay of gender with race and ethnicity in diagnosis and health care. Computational methods with big data analyses offer a unique opportunity to apply an intersectional lens to analyses, which can be difficult to accomplish with traditional survey methods. Intersectionality, a framework theorized by feminist legal scholar Kimberlé Crenshaw, help us to not only identify and address disparities, but can keep researchers from reproducing gender health disparities in our work (70). Public health scholar Lisa Bowleg has distilled the core tenets that are relevant to health research (71), which we apply to computational health research. Social identities like race, ethnicity, and gender are intersecting and multidimensional and should be examined accordingly. For example, the experience of Black women with health care should be examined specifically, rather than as the compounded experiences of white women + Black men. Analytic approaches using big data often allow for this level of specificity. As there are many social identities, intersectionality guides our research efforts to focus on the historically oppressed and marginalized peoples in order to achieve equity. Finally, rather than treating gender, race and ethnicity as individual predictive factors, we can shift our focus to understand how social identities interact with social systems embedded with racism and sexism. After all it is the interactions with social systems that produce the disparate health outcomes, but rather social identities themselves (71).

We should use big data to identify bias in diagnosis, treatment and outcomes among women, focusing on Black, Indigenous and Latina women. For example, large scale studies using EHR often have large enough sample sizes to conduct intersectional analyses of women with various racial and ethnic identities. Machine learning can also be used to develop models that perform better tan the standard of care for BIPOC women specifically, and in turn women broadly (66, 72). Computational approaches should be used to uncover and address bias in traditional standards of care.

There are some challenges. In a recent review of 164 studies that used EHR to develop machine learning models, 24% of studies did not report gender, and 64% did not report race/ethnicity (73). The computational health studies that include gender, typically do not report race and ethnicity data (73). When gender, race and ethnicity are excluded, it is impossible to identify gender disparities at large, and distinct outcomes among Black, Indigenous and Latina women, groups for whom we anticipate finding the largest disparities. Social identity omissions can also inadvertently worsen existing disparities. For example, while machine learning is based on language models that don’t include language used by Indigenous or Black women, those groups are systematically excluded from disease monitoring, and potentially from future allocations of resources and care.

These omissions can also introduce bias into machine learning models (75). (75) warn that even small amounts of bias in machine learning models can significantly impact the public’s health, as those biases are magnified across the population (75).

Measuring gender, race, and ethnicity is challenging, particularly when those identities are predicted using social media language or geolocation (74). However, there are best practices which are superior to excluding gender, race and ethnicity from computational health. When developed correctly, machine learning and AI modeling can help identify and develop automated workarounds for long held biases that drive many disparities, and ultimately to provide better care for all women (75, 66). We can learn from the emerging conversation about errors in the widespread misuse of race in medical corrections (76). There is also a research need dedicated to across the gender spec-trum including intersex, transgender and nonbinary individuals (77, 78, 79).

### 4.3. Limitations

This work is limited as we did not make a distinction between gender disparities driven by gender identity vs. those driven by biological sex difference. Further research is necessary in this area. As mentioned above, we also were unable to conduct a comprehensive search of the literature, although we will have identified key papers in this area indexed in MEDLINE, Embase and PsycINFO.

### 4.4. Conclusions

Big data holds many promises. The potential for a deeper understanding of what are the differences and what are the disparities between men and women could help improve health outcomes for patients. Therefore, for patients this could mean more appropriate care, for the clinician better evidence for making targeted decisions and for the policy maker the ability to make more equitable and effective policies. Understanding gender health disparities will help to inform decision-making, not at the individual patient level but also at the health policy level and help to provide a more equitable society.

## Data Availability

All data produced in the present work are contained in the manuscript

## ACKNOWLEDGMENTS

Dr. Stevens was supported in part by the National Institute on Drug Abuse (NIDA) of the National Institutes of Health under award number R21DA049572 - 01.

